# The incubation period for monkeypox cases confirmed in the Netherlands, May 2022

**DOI:** 10.1101/2022.06.09.22276068

**Authors:** Fuminari Miura, Catharina Else van Ewijk, Jantien A. Backer, Maria Xiridou, Eelco Franz, Eline Op de Coul, Diederik Brandwagt, Brigitte van Cleef, Gini van Rijckevorsel, Corien Swaan, Susan van den Hof, Jacco Wallinga

## Abstract

In May 2022 outbreaks of monkeypox have been reported in countries where the monkeypox virus is not endemic. We estimate the incubation period for monkeypox, using the reported time of exposure and symptom onset for 18 confirmed cases detected in the Netherlands up to 31st May 2022. The mean incubation period was 8.5 days, ranging from 4.2 to 17.3 days (5th to 95th percentiles). These findings underpin 21 days for monitoring or quarantining of case contacts.

## Background

Since the beginning of May 2022, monkeypox outbreaks have been reported in countries where the monkeypox virus is not endemic (1). Key public health measures to stop the spread of infection include active case finding, contact tracing, and quarantining close contacts. The incubation period of monkeypox has been reported to be up to 21 days, prompting public health institutes to recommend active monitoring and quarantine of close contacts for a minimum of 21 days after the last day of exposure (1–3).

The duration of the incubation period for monkeypox is known to depend on the transmission route (4). It is therefore essential to establish the distribution of the incubation period in the recent outbreaks. Whereas cases in previous outbreaks of monkeypox had traveled to endemic countries or had contact with infected animals (5), many cases in the current outbreaks have no documented history of travel to endemic countries and identify as men having sex with men (MSM) (6,7). High-risk contacts including sexual activity may play an important role in the current outbreak. Given these differences in exposure and route of transmission, the incubation period for monkeypox in the current outbreaks may also differ from values that have been reported before in settings with household or droplet transmission.

Here we report on the estimated incubation period of monkeypox using the reported time of exposure and symptom onset for confirmed monkeypox cases detected in the Netherlands up to 31st May 2022.

### Observed incubation periods

In the Netherlands, monkeypox was classified as a group A notifiable disease on 21 May 2022. As of 31 May, 31 monkeypox cases were laboratory-confirmed in the Netherlands. All cases were men and identified themselves as MSM, and the age range was 23–64 years old. At data collection, 18 cases had reported the symptom onset date and the most likely date of exposure as a single date or a limited number of consecutive dates, related to the attendance of an event where exposure was considered most likely.

We fitted parametric distributions to the observed incubation periods among 18 cases with symptom onset and exposure histories for monkeypox, using a likelihood-based approach, which allows for exposure to be a single time point or a time interval (8). The computation was implemented in R-4.0.5 (9) with a package {rstan}-2.21.2 (10). We compared three alternative parametric distributions: the lognormal, the gamma, and the Weibull distribution, and selected the best fitting distribution.

The reported incubation intervals for monkeypox were best described by a lognormal distribution (**Table 1**). Using this best-fitting distribution, the mean incubation period was estimated to be 8.5 days (95% credible intervals (CI): 6.6–10.9 days), with the 5th percentile of 4.2 days and the 95th percentile of 17.3 days (**Table 2**).

**Table 1.** Estimated mean of incubation period for different parametric distributions and computed goodness-of-fit.

|  | Mean (days) | WAIC* | LOOIC<sup>†</sup> |
| --- | --- | --- | --- |
| Lognormal | 8.5 [95%CI<sup>‡</sup>: 6.6–10.9] | 77.1 | 77.5 |
| Gamma | 9.1 [95%CI: 7.5–11.3] | 79.5 | 79.7 |
| Weibull | 9.6 [95%CI: 7.4–12.4] | 81.2 | 81.6 |

\*WAIC: Widely applicable information criterion. <sup>†</sup>LOOIC: Leave-One-Out Information Criterion; these values indicate the goodness-of-fit, where lower values indicate a better fit. <sup>‡</sup>CI, credible interval.

**Table 2.** Estimated percentiles of the incubation period for monkeypox, using different parametric distributions.

| Percentile | Lognormal |  | Gamma |  | Weibull |  |
| --- | --- | --- | --- | --- | --- | --- |
|  | Estimate | 95% CI* | Estimate | 95% CI | Estimate | 95% CI |
| 2.5<sup>th</sup> | 3.6 | 2.0–5.0 | 3.8 | 2.0–5.2 | 2.3 | 0.9–4.1 |
| 5<sup>th</sup> | 4.2 | 2.5–5.5 | 4.4 | 2.6–5.8 | 3.1 | 1.4–5.0 |
| 50<sup>th</sup> | 8.5 | 6.6–10.9 | 8.7 | 7.0–10.7 | 9.2 | 6.9–11.8 |
| 95<sup>th</sup> | 17.3 | 13.0–29.0 | 15.3 | 12.5–20.7 | 16.9 | 13.7–23.9 |
| 97.5<sup>th</sup> | 19.9 | 14.4–35.7 | 16.9 | 13.6–23.3 | 18.5 | 14.9–26.9 |
| 99<sup>th</sup> | 23.3 | 16.3–45.8 | 18.8 | 14.9–26.7 | 20.3 | 16.1–30.6 |

\*CI, credible interval.

Visual inspection revealed a good match between the fitted cumulative lognormal distribution function and the empirical cumulative distribution function, including the right tail of the distribution that describes the frequency of long incubation periods (**Figure 1**). The 2.5 percentile for the incubation period is estimated to be 3.6 days, and the 97.5 percentile is estimated to be 19.9 days (**Table 2**). An estimated two percent of all cases would develop first symptoms more than 21 days after being exposed.

**Fig 1.**
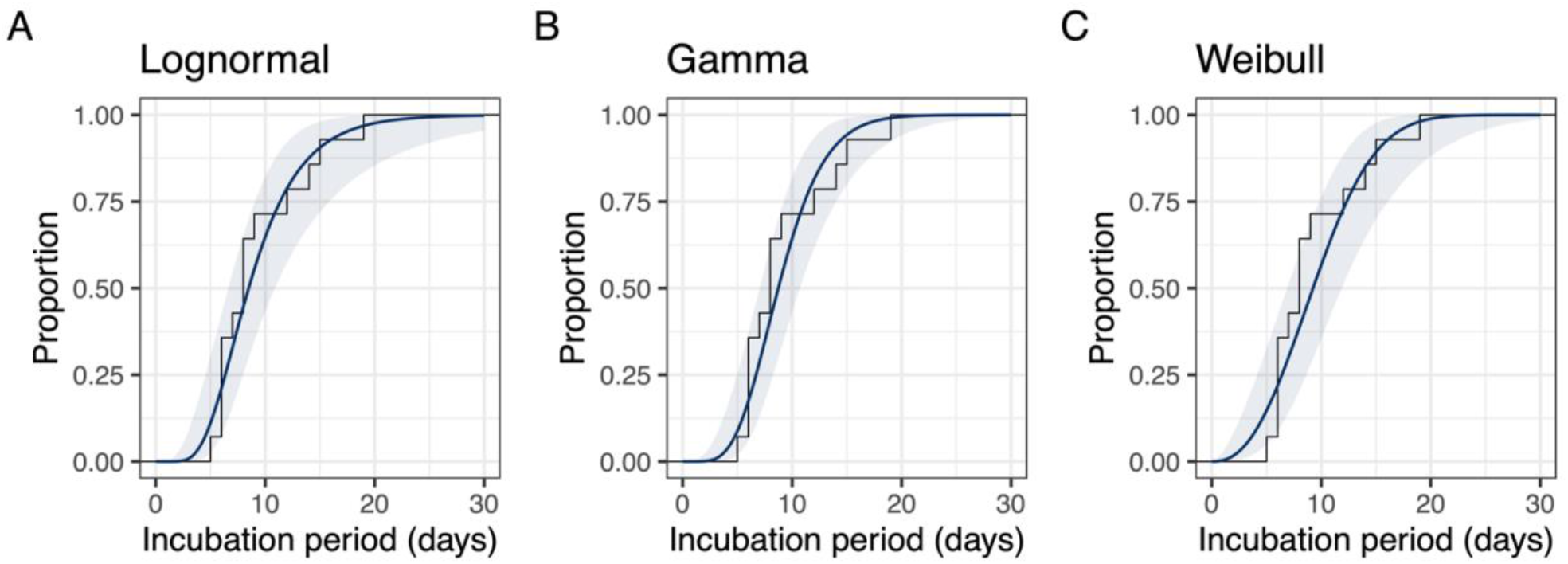
Estimated cumulative density function (colored line) and empirical cumulative density function (grey line) of incubation periods reported for monkeypox cases in the Netherlands.

## Discussion

This study provides empirical evidence for the distribution of the incubation period in a 2022 monkeypox outbreak, using the data on exposure histories and symptom onsets of monkeypox cases laboratory-confirmed by Polymerase Chain Reaction (PCR) in the Netherlands. The estimated 95th percentile of 17.3 days and the 97.5th percentile of 19.9 days can underpin the usage of 21 days for monitoring or quarantining close contacts of cases to limit further spread of the infection.

The duration of the incubation period has been reported to differ by route of transmission for monkeypox virus, smallpox, and vaccinia viruses (4). For non-invasive exposure (e.g. intact skin contact or droplet transmission) the typical incubation period of monkeypox is 13 days, and for complex and invasive exposures (e.g. contact with broken skin or mucous membranes), the typical incubation period is 9 days (4). These values are consistent with those of smallpox: about 12 days for outbreaks when exposure is predominant non-invasive (11,12) and about 9 days for inoculation when exposure is invasive (13). Our estimate of the mean incubation period of monkeypox in this outbreak of 8.5 days agrees is in line with the typical values for complex, invasive exposure. This result is supported by the epidemiological observation that all notified cases currently reported in the Netherlands are MSM with high-risk contact including sexual behavior. As most cases presented lesions in the anal and genital regions, direct skin contact or sexual transmission are the most likely routes of transmission among cases reported in the current outbreak.

If the reported incubation periods are those of the first observed cases in a growing outbreak, infected persons with a long incubation period would have a lower probability to be included, when compared with infected persons with a short incubation period. This suggests that the estimated incubation periods may suffer from downward bias, and that more than two percent of all infected cases would develop first symptoms more than 21 days after being exposed. In addition, the current estimate is based on 18 confirmed cases, and thus the continued monitoring of incubation periods of cases will provide more precision.

### Conclusion

In conclusion, this report presents a plausible range of incubation periods for the 2022 monkeypox outbreaks. The estimated mean incubation period is in line with previous findings for complex, invasive exposure to monkeypox. The estimated percentage of monkeypox cases that would develop symptoms after the conclusion of 21 days period is approximately two percent. These findings suffice for the current use of 21 days for quarantining, but as the outbreaks grow and cases can be infected by different transmission routes, continued monitoring of the incubation period for monkeypox is necessary.

## Data Availability

Anonymized data and all codes used for analysis and visualization are available on Github (https://github.com/fmiura/MpxInc_2022).

## Acknowledgement

We thank the Public Health Services for their effort to collect the epidemiological data.

## Author contributions

Conceptualization: FM JW.

Data curation: FM CvE EF EdC DB BvC GvR CS.

Formal analysis: FM JW.

Investigation: FM CvE JW.

Methodology: FM JB JW.

Software: FM.

Validation: FM.

Visualization: FM.

Writing – original draft: FM JW.

Writing – review & editing: FM CvE JB MX EF EdC DB BvC GvR CS SvdF JW.

